# Heterogeneous Mental Health Development During the COVID-19 Pandemic in the United Kingdom

**DOI:** 10.1101/2021.02.24.21251565

**Authors:** Lea Ellwardt, Patrick Präg

## Abstract

The COVID-19 pandemic and the mitigation measures by governments have upended the economic and social lives of many, leading to widespread psychological distress. We explore heterogeneity in trajectories of psychological distress during the pandemic in the United Kingdom and relate this heterogeneity to socio-demographic and health factors. We analyze nine waves of longitudinal, nationally representative survey data from the UK Household Longitudinal Study (*N* =15,914), covering the period from early 2020 to mid-2021. First, latent class mixture modelling (LCCM) is used to identify trajectories of psychological distress. Second, associations of the trajectories with covariates are tested with multinomial logistic regressions. We find four different trajectories of distress: continuously low, temporarily elevated, repeatedly elevated, and continuously elevated distress. Nearly two fifths of the population experienced severely elevated risks of distress during the pandemic. Long-term distress was highest among younger people, women, people living without a partner, those who had no work or lost income, and those with previous health conditions or COVID-19 symptoms. Given the threat of persistent stress on health, policy measures should be sensitized to the unintended yet far-reaching consequences of non-pharmaceutical interventions.

## Introduction

The COVID-19 pandemic has challenged the routines of everyday life on an unprecedented scale and for many caused worries about aspects such as threats to physical health and financial security, along with the distressing experience of social isolation. However, there is considerable variation in the prevalence of psychological distress (Fancourt *et al*., 2021; Holmes *et al*., 2020; Niedzwiedz *et al*., 2021a; O’Connor *et al*., 2021; Pierce *et al*., 2020, 2021; McGinty *et al*., 2020; Giuntella *et al*., 2021; Buyukkececi, 2021; Van Winkle *et al*., forthcoming), both across social groups and across countries.

During the pandemic, average mental health deteriorated for the whole population, and research has examined variation in this deterioration across subpopulations (Niedzwiedz *et al*., 2021a; Proto and Quintana-Domeque, 2021; Chandola *et al*., 2021). These subpopulations are typically segmented ex ante in line with theoretical expectations by discrete socio-demographic characteristics, such as social class, ethnicity, or gender. We argue that the development of the individuals’ mental health during the COVID-19 pandemic was heterogeneous. Further, we argue that much of the sociodemographic predictors examined in previous research of this heterogeneity might not be as deterministic as that research assumes: Several distinct developments are feasible that do not unanimously pertain to specific sociodemographic subpopulations and thereby remain hidden in explanatory analyses. A recent exception is the study by Pierce *et al*. (2021), who modelled trajectories during the first lockdown in the United Kingdom with a latent class analysis. We use similar data-driven exploration, in which we compare and classify individuals based on their mental health development over time, and inspect socio-demographic profiles ex post. The aim of the present study was, first, to detect the number and prevalences of psychological distress trajectories over the COVID-19 pandemic, and second, to relate these trajectories to individuals’ socio-demographic and health characteristics.

People vary both in their susceptibility to pandemic-induced stressors (Chandola *et al*., 2021; Kuhn *et al*., 2021; Molteni *et al*., 2021; Shen and Bartram, 2021) and their ability to successfully cope with them. This results in different trajectories of psychological distress. For example, while some individuals suffer from increasing psychological distress from the onset (linear growth), others are affected at first and distress levels off again later (inverted u-curve), and others remain resilient to the external stressor altogether. Similarly, the economic recession associated with the pandemic was v-shaped for high-wage workers, and was much deeper and ongoing for low-wage workers (Chetty *et al*., 2020; Holst *et al*., 2021; Witteveen and Velthorst, 2020). We expect mental health to follow heterogeneous developments which are associated with individual characteristics.

The British government announced on March 23, 2020 that residents must stay at home and some businesses have to close, one of the longest and harshest government measures of the pandemic. This lockdown was gradually eased until July 4 when most businesses were allowed to open again. From late July onwards, a series of limited and often local restrictions was put in place. From November 5, England went into a second, four-week long lockdown, which differed from the first in that schools and universities remained open, and from December onwards returned to a system of local restrictions. In early January 2021, the United Kingdom went into a third lockdown that ended gradually: in March, schools reopened, in April, individuals in groups of up to six were allowed to meet again outdoors, outdoor pubs, restaurants and non-essential shops reopened in England, in mid-May 2021, outdoor social contact became unregulated again, individuals in groups of up to six were allowed to meet indoors, and restaurants and hotels could re-open.

Ignoring the existence of simultaneous yet distinct trajectories in coping with the lockdown likely obscures important differences across subpopulations. Previous research has shown, for example, that disadvantaged social groups are more vulnerable to stressors, have worse mental and physical health, and are less resilient to adverse life events than advantaged groups (Thoits, 2010). Likewise, the unique case of repeated lockdowns imposed increased informal care responsibilities on women (Fodor *et al*., 2021; Möhring *et al*., 2021; Schmid *et al*., 2021; Zoch *et al*., 2021) and heightened risks of loneliness for people living alone (van Tilburg *et al*., 2021; Elmer *et al*., 2020).

Trajectories were retrieved from a time series of mental health data covering the period before, during, and after the three lockdowns in the United Kingdom. In our analyses we, first, employed latent class mixture modelling (LCCM) (Collins and Lanza, 2010) to estimate trajectories of change in psychological distress. This data-reduction technique classifies individuals within a population and probabilistically assigns them into latent classes. Each class represents a distinct subpopulation of individuals with highly similar trajectories. Second, using multinomial logistic regression, we examined factors associated with the different trajectories.

Our study extends a recent study (Pierce *et al*., 2021) with a similar analytical approach of the same data and similar findings by extending the observation window. Pierce *et al*. (2021) restricted their analyses to the period until September 2020, we add three additional waves of data until May 2021.

## Results

Development of psychological distress followed four distinct trajectories, as revealed by the latent class mixture models. The model fit levelled off for greater numbers of trajectories, i.e. the log-likelihood based statistics (van der Nest *et al*., 2020) improved only slightly in the five-trajectory solution, as seen in Table 1. The five-trajectory solution was still well-fitted, but generated two highly similar trajectories in the first half of the observation window, and three highly similar trajectories in the second half. The model with four trajectories was therefore more parsimonious and expedient in its further interpretation.

**Table 1:** Fit statistics and class prevalence for models with *k* latent trajectories

| No. trajectories ($k$) | BIC | AIC | LL | % Trajectory 1 | % Trajectory 2 | % Trajectory 3 | % Trajectory 4 | % Trajectory 5 |
| --- | --- | --- | --- | --- | --- | --- | --- | --- |
| 1 | -64339.4 | -64324.1 | -64320.1 | 100 |  |  |  |  |
| 2 | -50974.6 | -50951.6 | -50945.6 | 70 | 30 |  |  |  |
| 3 | -49710.9 | -49668.7 | -49657.7 | 57 | 29 | 14 |  |  |
| 4 | -49355.2 | -49293.8 | -49277.8 | 53 | 8 | 24 | 15 |  |
| 5 | -49250.3 | -49169.7 | -49148.7 | 26 | 36 | 21 | 4 | 13 |

*Notes:* Based on individuals with at least three completed waves on psychological distress ( $N = 15,914$ ). BIC: Bayes Information Criterion, AIC: Akaike Information Criterion, LL: Log-likelihood.

The four psychological distress trajectories are visualized in Figure 1. Continuously low (trajectory 1): The largest group included half of the participants (53.2%), whose likelihood of psychological distress remained permanently close to zero. Temporarily elevated (trajectory 2): The smallest group contained 8.0% of the participants, who started out with a rather low likelihood of distress, comparable to those in trajectory 1. Crucially, risk of psychological distress rapidly rose to two thirds at the beginning of the pandemic. This stark elevation was of temporal nature, as distress recovered quickly back to zero after the first lockdown and remained stably low thereafter. Repeatedly elevated (trajectory 3): The second-largest group comprised one fourth of the participants (24.0%). Their likelihood of psychological distress was roughly one third at the start and peaked during all periods of lockdown, reaching its maximum of one half in the third lockdown.

**Figure 1:**
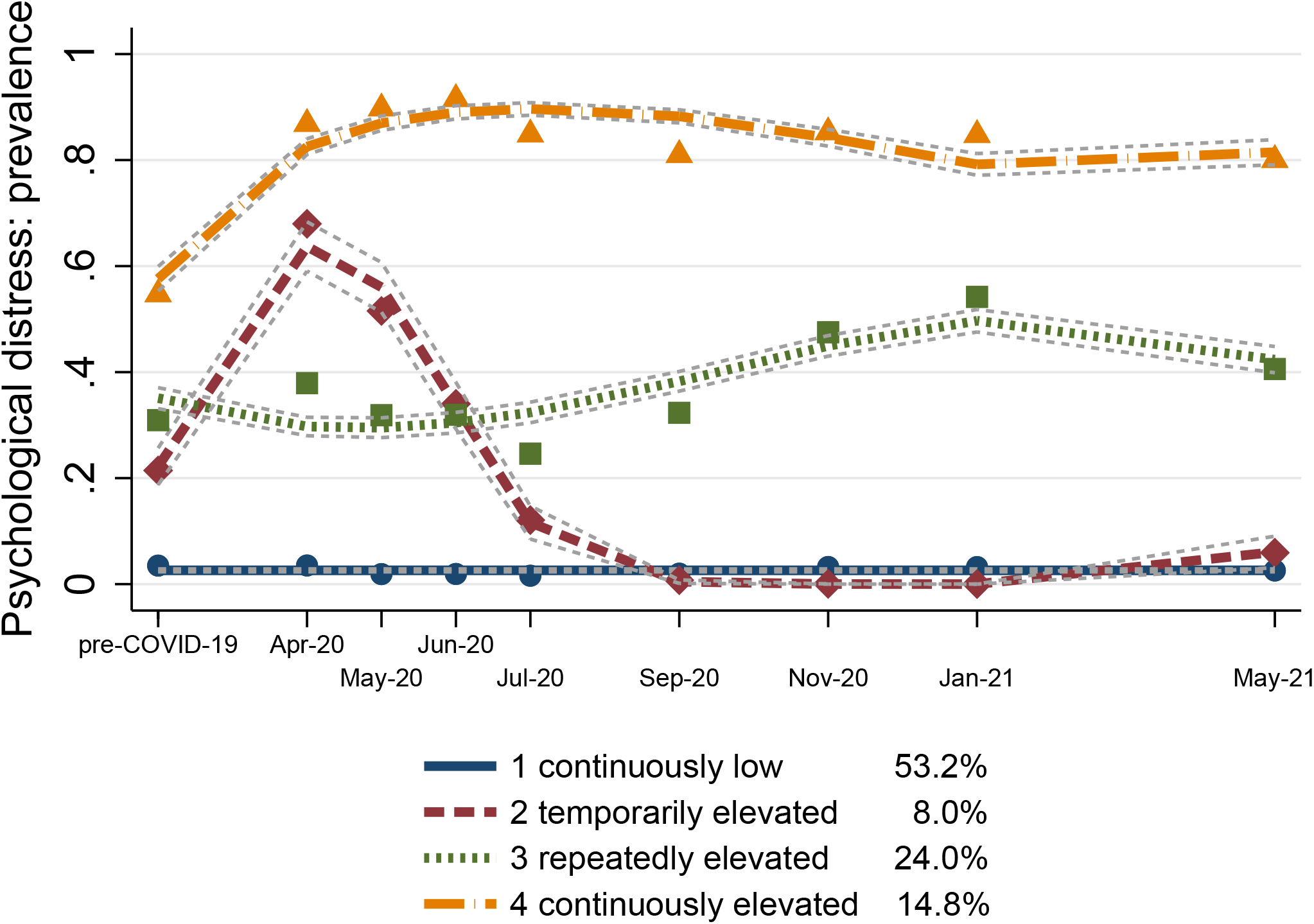
Four latent trajectories of psychological distress. Error bounds represent 95% confidence intervals. Based on Understanding Society COVID-19 Study (*N* = 15,914).

Continuously elevated (trajectory 4): The third-largest group comprised of 14.8% of the participants, who had a consistently high likelihood of psychological distress, with the vast majority of the individuals reporting distress at all time points. Yet, also in this trajectory, mental health covaried with the start of the pandemic. The likelihood of distress reached a near-maximum during the first lockdown and slightly declined thereafter, however, without returning to its initial level. Taking trajectories 3 and 4 together, about two fifths of the population experienced a significant and severe elevation of psychological distress during the whole observation period.

To examine whether socio-demographic and health variables are associated with being on the four trajectories, individuals were discretely assigned to the latent trajectory with the highest posterior probability. Higher means of this probability in a trajectory indicate greater certainty in the assignment procedure. Mean probabilities ranged from .69 for trajectory 2 to .89 for trajectory 1. Table 2 presents the distribution of the covariates across trajectories. The assigned trajectories served as the outcome in a multinomial logistic regression model. The association of the covariates with each of the four trajectories are expressed as average marginal effects (AME’s) shown in Figure 2. AME’s reflect the average change in a trajectory’s probability when a covariate increases by one unit.

**Table 2:** Proportions/means of covariates by trajectories

| Covariate | Trajectory 1<br>Continuously<br>low | Trajectory 2<br>Temporarily<br>elevated | Trajectory 3<br>Repeatedly<br>elevated | Trajectory 4<br>Continuously<br>elevated | Chi-square test<br>*p*-value |
| --- | --- | --- | --- | --- | --- |
| Age groups: |  |  |  |  | < 0.0001 |
| 24 years and younger | 0.03 | 0.06 | 0.05 | 0.06 | . |
| 25–44 years | 0.21 | 0.34 | 0.31 | 0.31 | . |
| 45–64 years | 0.45 | 0.45 | 0.43 | 0.46 | . |
| 65 years and older | 0.31 | 0.15 | 0.20 | 0.17 | . |
| Female (*ref.* male) | 0.51 | 0.67 | 0.68 | 0.69 | < 0.0001 |
| Non-white (*ref.* white) | 0.10 | 0.12 | 0.10 | 0.13 | < 0.0001 |
| Living with partner | 0.78 | 0.71 | 0.71 | 0.63 | < 0.0001 |
| Child(ren) in household | 0.29 | 0.39 | 0.37 | 0.35 | < 0.0001 |
| Single parent | 0.04 | 0.06 | 0.07 | 0.09 | < 0.0001 |
| Worked pre-COVID | 0.63 | 0.75 | 0.70 | 0.65 | < 0.0001 |
| Reported COVID-19 symptoms | 0.19 | 0.29 | 0.28 | 0.33 | < 0.0001 |
| No health conditions | 0.51 | 0.56 | 0.48 | 0.41 | < 0.0001 |
| Social class: |  |  |  |  | 0.294 |
| Working class | 0.27 | 0.26 | 0.25 | 0.28 | . |
| Intermediate | 0.24 | 0.23 | 0.24 | 0.24 | . |
| Professional | 0.49 | 0.51 | 0.50 | 0.48 | . |
| Earnings pre-COVID (log)<sup>a</sup> | 7.26 | 7.37 | 7.34 | 7.23 | < 0.0001 |
| Lost earnings | 0.34 | 0.42 | 0.37 | 0.36 | < 0.0001 |
| *N* individuals | 8,075 | 741 | 3,113 | 1,991 | 13,920 |

*Notes:* Based on individuals with complete information on all covariates at baseline. <sup>a</sup> Mean-imputed.

**Figure 2:**
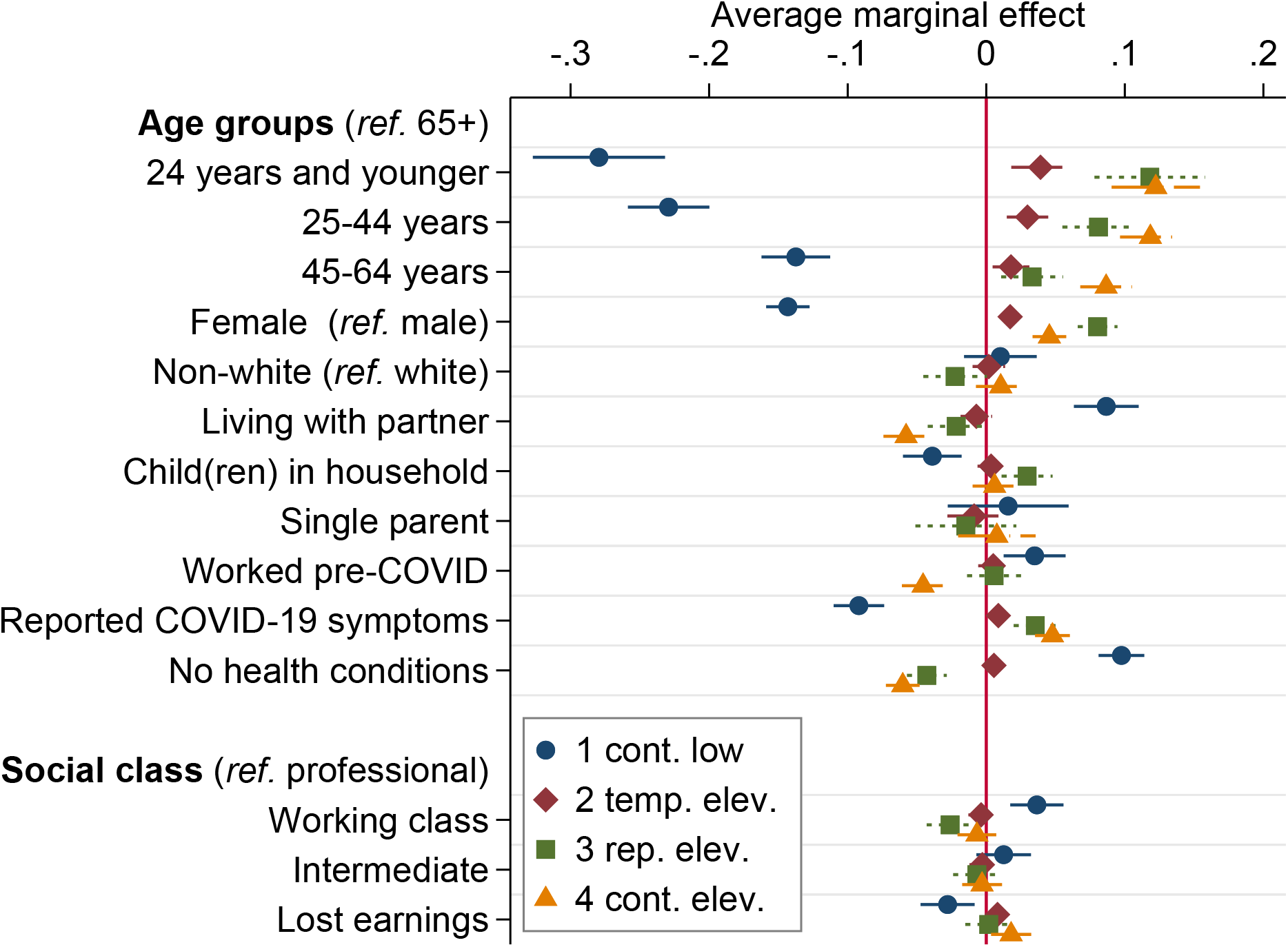
Average marginal effects (AME’s) of multinomial regression model of trajectory membership. Notes: Average change in a trajectory’s probability when a covariate increases by one unit, from multinomial model with the trajectories of psychological distress as the outcome: (1) continuously low, (2) temporarily elevated, (3) repeatedly elevated, (4) continuously elevated. Error bars represent 95% confidence intervals. AME’s for region, household earnings, and missing earnings not shown. Based on Understanding Society COVID-19 Study (*N* = 13,920).

The following focuses on changes in mental health, thus on individuals with elevated stress levels in trajectories 2 to 4. The risk of experiencing any of these trajectories was highest for young and female individuals and those reporting COVID-19 symptoms. Repeated distress was more probable for people living with children in the household. Furthermore, the risk for continued distress was highest for individuals who lived without a partner, had not worked before the pandemic, mentioned pre-pandemic health conditions, and lost substantial income during the course of the pandemic. For the latter sub-populations, the initial probability of 14.8% in trajectory 4 shifted upwards by another 1.8 to 12.2 percentage points (e.g., income loss was associated with a total risk of 16.6% and being aged 24 years or younger had an adjusted risk of 27.0%). This implied a strong impact of some of these covariates on developing continuously elevated psychological distress.

## Discussion

The unparalleled shock of the COVID-19 pandemic has provoked a natural stress test for entire societies. We took a perspective similar to those of policy makers and stakeholders by describing which trajectories of psychological distress occurred in the United Kingdom. We next described the most affected subpopulation in terms of socio-demographic profiles, rather than single out presumably vulnerable subpopulations for inspection *a priori*.

Our results replicate and extend the findings by Pierce *et al*. (2021), who used a similar model and the same data to study the first lockdown in the UK. Their findings revealed comparable trajectories to ours, with one main nuance: The continuously low-distress group was refined into two subgroups (‘consistently good’ and ‘consistently very good’ mental health), because the authors used a mean GHQ-score instead of a dichotomous cut-off for distress. In contrast to Proto and Quintana-Domeque (2021), we did not find an average increase in psychological distress for the whole UK sample. However, their study covered changes between pre-COVID-19 and April 2020 only, i.e. the first two measurements in our study design. This snapshot could not capture short-term hiccups in distress that bounced back after the lockdown, as identified in trajectory 2. But even though in our study the majority either proved resilient or recovered quickly, two in five individuals suffered from psychological distress until after the observation period. This vulnerable group was not solely defined by the usual risk factors of poor health. These findings closely resemble previous findings from the United Kingdom, where one out of four individuals reported depressive symptoms and one out of five individuals suffered from anxiety (O’Connor *et al*., 2021; Pierce *et al*., 2021). Using the same data as in our study and group-based latent growth mixture models on depressive symptoms, an outcome closely related to our measure of psychological distress, Iob *et al*. (2020) found three latent trajectories. These included low (60%), moderate (29%) and severe (11%) depressive symptoms during lockdown.

As in previous research, in our study risks were greater for those who are younger, female (Banks and Xu, 2020; Pierce *et al*., 2021), individuals living without a partner (Fancourt *et al*., 2021; Pierce *et al*., 2020), individuals with COVID-19 related symptoms (Chandola *et al*., 2021; Li and Wang, 2020), and those who lost income (Bu *et al*., 2020b,a; Wright *et al*., 2021). Crucially, individuals who fail to buffer the long-term exposure of distress are vulnerable to a variety of negative health outcomes, including poor physical health, morbidity, and mortality (Barry *et al*., 2020; Thoits, 2010). More dramatically, chronic psychological stress has been related to lower immunity and, as a result, higher susceptibility of the common cold, influenza, infectious diseases and upper respiratory illness (Cohen *et al*., 1991). This means that measures aimed at mitigating the COVID-19 pandemic might promote risk factors for catching the coronavirus, if these measures trigger stress among the broader public, and thereby backfire on a large scale. Distress might at least partly offset beneficial health consequences of governmental measures (Mader and Rüttenauer, 2021). Reducing distress should therefore be key in policy-making aiming to safeguard public health, and even more so during repeated lockdowns that accumulate stressors into constant emotional exhaustion. In our analysis, particularly women and those living with children were prone to a peak of distress during the third lockdown. Mounting challenges of orchestrating family care and homeschooling duties likely helped pave this way.

A limitation of the present study was the use of self-reported psychological distress. The pandemic is still ongoing, and the months and years to come will show whether deterioration in mental health manifests into heightened therapeutic need and psychiatric intervention, or whether societies put this collective trauma behind. Moreover, the development of psychological distress may be investigated in combination with correlated health variables, such as drinking behavior, through modelling joint trajectories.

The heterogeneity in the development of mental health during the pandemic emphasizes peoples’ idiosyncrasy in responses to extreme changes. Likewise, policy-makers will need to be careful in identifying and supporting groups at risk. Addressed health risks should not merely prioritize physical outcomes directly related to the COVID-19 disease, but include longer-term consequences in the domain of mental and social well-being of the general public (Holmes *et al*., 2020).

## Methods

We analyze data from the Understanding Society COVID-19 Study (University of Essex and Institute for Social and Economic Research, 2021; Platt *et al*., 2020). The UK Household Longitudinal Study “Understanding Society” is a long-running, nationally representative panel survey that annually interviews those members 16 years or older of participating households. During the COVID-19 pandemic, additional monthly online and phone interviews were conducted in April, May, June, July, September, and November 2020 as well as in January and May 2021. The University of Essex Ethics Committee approved the data collection and informed consent was obtained from all participants. No ethics approval was necessary for this secondary data analysis. All experiments were performed in accordance with relevant guidelines and regulations. The analytical sample is restricted to 15,914 participants with information on psychological distress for a minimum of three waves. This is to increase validity in the modelling of the participants’ trajectories.

Psychological distress is measured with the General Health Questionnaire (GHQ-12, Goldberg, 1972). Participants are presented with twelve questions (e.g. “Have you recently been feeling unhappy or depressed?”), and those who respond “Much more than usual” or “Rather more than usual” receive a score of 1, while those responding “No more than usual” or “Not at all” receive a score of 0 (the so-called “caseness” scoring). Summated scores of four or more (out of twelve) are usually considered as distressed. As done in previous research (Niedzwiedz *et al*., 2021b), we dichotomized scores accordingly into presence (1) versus absence (0) of psychological distress in every of the nine measurement waves.

Covariates included dummy variables for the countries Scotland, Wales, and Northern Ireland (England was the reference category), age (24 years and younger, 25–44 years, 45–64 years, 65 years and older), gender (female = 1 vs. male = 0), race (Non-White = 1 vs. White = 0), and living with partner (yes = 1 vs. no = 0). Further, social class was accounted for based on a collapsed version of the NS-SEC social class scheme (working class, intermediate, professional) (Rose and Pevalin, 2003), an established measure of social class in the United Kingdom (Präg and Richards, 2019) and a known predictor of mental health (Gugushvili and Präg, 2021; Präg and Gugushvili, 2021). Monthly earnings (log-transformed and mean-imputed), a dummy variable indicating an income loss of 15% or more during the pandemic, and two dummy variables to adjust for missing values in the previous earnings variables were also included in the models. Other socio-demographic covariates regarded sharing the household with one or more children aged under 19 years, being a single parent (composed from the aforementioned partner and child variables), being employed or self-employed (all: yes = 1 vs. no = 1). Health covariates concerned having had symptoms of COVID-19 and mentioning any diagnosed health conditions (both: yes = 1 vs. no = 0). All covariates stemmed from baseline of the COVID-19 study module, April 2020.

Because we had no *a priori* expectations of specific distributions, we opted for an explorative approach to determine number and prevalence of trajectories, using the nine repeated measurements of psychological distress. Mixture models for the clustering of longitudinal data series identify latent subpopulations that share similar trajectories (van der Nest *et al*., 2020). These trajectories, which are highly comparable within subpopulations, are deemed mutually exclusive between subpopulations. To determine the number of distinct trajectories, we first estimated a set of models for a varying number of *k* trajectories, starting with *k* = 1, and sequentially increasing *k* by one, until the model fit leveled off or deteriorated.

Second, we proceeded with the model with the best fit to investigate associations of covariates with each of the *k* trajectories. Specifically, a participant’s trajectory with the highest probability served as the categorical outcome in a multinomial logit model. The sample in this model was reduced to participants with complete covariates. We show average marginal effects (AME’s), which reflect the average change in a trajectory’s probability when a covariate increases by one unit. The statistical analysis was performed in Stata 17 using the command ‘traj’ (version May 17, 2020) for estimating group-based trajectory models (Jones and Nagin, 2013).

## Data Availability

The data analyzed in this study are publicly available (University of Essex and Institute for Social and Economic Research, 2021) and a replication package can be found on-line (Ellwardt and Praeg, 2021).

https://dx.doi.org/10.17605/OSF.IO/ZXQE6

## Data availability

The data analyzed in this study are publicly available from the UK Data Service (University of Essex and Institute for Social and Economic Research, 2021) to registered users and a replication package for all analyses shown is available online (Ellwardt and Präg, 2021b).

## Author contributions statement

L.E. and P.P. conceptualized the study, executed the analysis, and wrote the manuscript.

## Additional information

The authors of this study declare no competing interests.

## References

Banks, James and Xiaowei Xu, 2020. ‘The Mental Health Effects of the First Two Months of Lockdown during the COVID-19 Pandemic in the UK.’ Fiscal Studies 41(3): 685–708. doi: 10.1111/1475-5890.12239.

Barry, Vaughn, Mary E. Stout, Mary Ellen Lynch, Shanna Mattis, Duc Q. Tran, Ana Antun, Maria J. A. Ribeiro, Sidney F. Stein, and Christine L. Kempton, 2020. ‘The Effect of Psychological Distress on Health Outcomes: A Systematic Review and Meta-Analysis of Prospective Studies.’ Journal of Health Psychology 25(2): 227–239. doi: 10.1177/1359105319842931.

Bu, Feifei, Andrew Steptoe, and Daisy Fancourt, 2020a. ‘Loneliness During a Strict Lockdown: Trajectories and Predictors During the COVID-19 Pandemic in 38,217 United Kingdom Adults.’ Social Science and Medicine 265(113521): 1–6. doi: 10.1016/j.socscimed.2020.113521.

Bu, Feifei, Andrew Steptoe, and Daisy Fancourt, 2020b. ‘Who Is Lonely in Lockdown? Cross-Cohort Analyses of Predictors of Loneliness Before and During the COVID-19 Pandemic.’ Public Health 186: 31–34. doi: 10.1016/j.puhe.2020.06.036.

Buyukkececi, Zafer, 2021. ‘Cross-Country Differences in Anxiety and Behavioral Response to the COVID-19 Pandemic.’ European Societies 23(S1): S417–S447. doi: 10.1080/14616696.2020.1828975.

Chandola, Tarani, Meena Kumari, Cara L. Booker, and Michaela Benzeval, 2021. ‘The Mental Health Impact of COVID-19 and Lockdown-Related Stressors among Adults in the UK.’ Psychological Medicine pp. 1–10. doi:10.1017/S0033291720005048.

Chetty, Raj, John N. Friedman, Nathaniel Hendren, Michael Stepner, and The Opportunity Insights Team, 2020. ‘The Economic Impacts of COVID-19: Evidence from a New Public Database Built Using Private Sector Data.’ National Bureau of Economic Research Working Paper Series 27431: 1–64. doi: 10.3386/w27431.

Cohen, Sheldon, David A. J. Tyrrell, and Andrew P. Smith, 1991. ‘Psychological Stress and Susceptibility to the Common Cold.’ New England Journal of Medicine 325(9): 606–612. doi: 10.1056/NEJM199108293250903.

Collins, Linda M. and Stephanie T. Lanza, 2010. Latent Class and Latent Transition Analysis. With Applications in the Social, Behavioral, and Health Sciences. New York: Wiley. doi: 10.1002/9780470567333.

Ellwardt, Lea and Patrick Präg, 2021a. ‘Heterogeneous Mental Health Development During the COVID-19 Pandemic in the United Kingdom.’ CREST Working Paper Series 2021–07.

Ellwardt, Lea and Patrick Präg, 2021b. ‘Replication Materials to: Heterogeneous Mental Health Development During the COVID-19 Pandemic in the United Kingdom.’ Open Science Framework doi: 10.17605/OSF.IO/ZXQE6.

Elmer, Timon, Kieran Mepham, and Christoph Stadtfeld, 2020. ‘Students under Lockdown: Comparisons of Students’ Social Networks and Mental Health Before and During the COVID-19 Crisis in Switzerland.’ Plos One 15(7): e0236337. doi: 10.1371/journal.pone.0236337.

Fancourt, Daisy, Andrew Steptoe, and Feifei Bu, 2021. ‘Trajectories of Anxiety and Depressive Symptoms During Enforced Isolation Due to COVID-19 in England: A Longitudinal Observational Study.’ Lancet Psychiatry 8(2): 141–149. doi: 10.1016/S2215-0366(20)30482-X.

Fodor, Eva, Anikó Gregor, Júlia Koltai, and Eszter Kováts, 2021. ‘The Impact of COVID-19 on the Gender Division of Childcare Work in Hungary.’ European Societies 23(S1): S95–S110. doi:10.1080/14616696.2020.1817522.

Giuntella, Osea, Kelly Hyde, Silvia Saccardo, and Sally Sadoff, 2021. ‘Lifestyle and Mental Health Disruptions During COVID-19.’ Proceedings of the National Academy of Sciences 118(9): e2016632118. doi:10.1073/pnas.2016632118.

Goldberg, David P., 1972. The Detection of Psychiatric Illness by Questionnaire. Oxford: Oxford University Press.

Gugushvili, Alexi and Patrick Präg, 2021. ‘Intergenerational Social Mobility and Health in Russia: Mind over Matter?’ Advances in Life Course Research 47(100390): 1–13. doi: 10.1016/j.alcr.2020.100390.

Holmes, Emily A., Rory C. O’Connor, V. Hugh Perry, Irene Tracey, Simon Wessely, Louise Arseneault, Clive Ballard, Helen Christensen, Roxane Cohen Silver, Ian Everall, Tamsin Ford, Ann John, Thomas Kabir, Kate King, Ira Madan, Susan Michie, Andrew K. Przybylski, Roz Shafran, Angela Sweeney, Carol M. Worthman, Lucy Yardley, Katherine Cowan, Claire Cope, Matthew Hotopf, and Ed Bullmore, 2020. ‘Multidisciplinary Research Priorities for the COVID-19 Pandemic: A Call for Action for Mental Health Science.’ Lancet Psychiatry 7(6): 547–560. doi: 10.1016/S2215-0366(20)30168-1.

Holst, Hajo, Agnes Fessler, and Steffen Niehoff, 2021. ‘COVID-19, Social Class, and Work Experience in Germany: Inequalities in Work-Related Health and Economic Risks.’ European Societies 23(S1): S495–S512. doi:10.1080/14616696.2020.1828979.

Iob, Eleonora, Philipp Frank, Andrew Steptoe, and Daisy Fancourt, 2020. ‘Levels of Severity of Depressive Symptoms Among At-Risk Groups in the UK During the COVID-19 Pandemic.’ JAMA Network Open 3(10): e2026064–e2026064. doi: 10.1001/jamanetworkopen.2020.26064.

Jones, Bobby L. and Daniel S. Nagin, 2013. ‘A Note on a Stata Plugin for Estimating Group-Based Trajectory Models.’ Sociological Methods & Research 42(4): 608–613. doi: 10.1177/0049124113503141.

Kuhn, Ursina, Hannah S. Klaas, Erika Antal, Nora Dasoki, Florence Lebert, Oliver Lipps, Gian-Andrea Monsch, Jan-Erik Refle, Valérie-Anne Ryser, Robin Tillmann, and Marieke Voorpostel, 2021. ‘Who Is Most Affected by the Corona Crisis? An Analysis of Changes in Stress and Wellbeing in Switzerland.’ European Societies 23(S1): S942–S956. doi:10.1080/14616696.2020.1839671.

Li, Lambert Zixin and Senhu Wang, 2020. ‘Prevalence and Predictors of General Psychiatric Disorders and Loneliness During COVID-19 in the United Kingdom.’ Psychiatry Research 291(113267): 1–6. doi:10.1016/j.psychres.2020.113267.

Mader, Sebastian and Tobias Rüttenauer, 2021. ‘The Effects of Nonpharmaceutical Interventions on COVID-19-Related Mortality: A Generalized Synthetic Control Approach Across 169 countries.’ SocArXiv doi:10.31235/osf.io/v2ef8.

McGinty, Emma E., Rachel Presskreischer, Hahrie Han, and Colleen L. Barry, 2020. ‘Psychological Distress and Loneliness Reported by US Adults in 2018 and April 2020.’ JAMA 324(1): 93–94. doi:10.1001/jama.2020.9740.

Möhring, Katja, Elias Naumann, Maximiliane Reifenscheid, Alexander Wenz, Tobias Rettig, Ulrich Krieger, Sabine Friedel, Marina Finkel, Carina Cornesse, and Annelies G. Blom, 2021. ‘The COVID-19 Pandemic and Subjective Wellbeing: Longitudinal Evidence on Satisfaction With Work and Family.’ European Societies 23(S1): S601–S617. doi:10.1080/14616696.2020.1833066.

Molteni, Francesco, Riccardo Ladini, Ferruccio Biolcati, Antonio M. Chiesi, Giulia Maria Dotti Sani, Simona Guglielmi, Marco Maraffi, Andrea Pedrazzani, Paolo Segatti, and Cristiano Vezzoni, 2021. ‘Searching for Comfort in Religion: Insecurity and Religious Behavior During the COVID19 Pandemic in Italy.’ European Societies 23(S1): S704–S720. doi:10.1080/14616696.2020.1836383.

Niedzwiedz, Claire L., Michaela Benzeval, Kirsten Hainey, Alastair H. Leyland, and Srinivasa Vittal Katikireddi, 2021a. ‘Psychological Distress Among People with Probable COVID-19 Infection: Analysis of the UK Household Longitudinal Study.’ BJPsych Open 7(3): 1–3. doi:10.1192/bjo.2021.63.

Niedzwiedz, Claire L., Michael James Green, Michaela Benzeval, Desmond Campbell, Peter Craig, Evangelia Demou, Alastair Leyland, Anna Pearce, Rachel Thomson, Elise Whitley, and Srinivasa Vittal Katikireddi, 2021b. ‘Mental Health and Health Behaviors Before and During the Initial Phase of the COVID-19 Lockdown: Longitudinal Analyses of the UK Household Longitudinal Study.’ Journal of Epidemiology and Community Health 75(3): 224–231. doi: 10.1136/jech-2020-215060.

O’Connor Rory C., Karen Wetherall, Seonaid Cleare, Heather McClelland, Ambrose J. Melson, Claire L. Niedzwiedz, Ronan E. O’Carroll, Daryl B. O’Connor, Steve Platt, Elizabeth Scowcroft, Billy Watson, Tiago Zortea, Eamonn Ferguson, and Kathryn A. Robb, 2021. ‘Mental Health and Well-being During the COVID-19 Pandemic: Longitudinal Analyses of Adults in the UK COVID-19 Mental Health and Wellbeing Study.’ British Journal of Psychiatry doi: 10.1192/bjp.2020.212.

Pierce, Matthias, Holly Hope, Tamsin Ford, Stephani Hatch, Matthew Hotopf, Ann John, Evangelos Kontopantelis, Roger Webb, Simon Wessely, Sally McManus, and Kathryn M. Abel, 2020. ‘Mental Health Before and During the COVID-19 Pandemic: A Longitudinal Probability Sample Survey of the UK Population.’ Lancet Psychiatry 7(10): 883–892. doi:10.1016/S2215-0366(20)30308-4.

Pierce, Matthias, Sally McManus, Holly Hope, Matthew Hotopf, Tamsin Ford, Stephani L. Hatch, Ann John, Evangelos Kontopantelis, Roger T. Webb, Simon Wessely, and Kathryn M. Abel, 2021. ‘Mental Health Responses to the COVID-19 Pandemic: A Latent Class Trajectory Analysis Using Longitudinal UK Data.’ Lancet Psychiatry 8(7): 610–619. doi:10.1016/S2215-0366(21)00151-6.

Platt, Lucinda, Gundi Knies, Renee Luthra, Alita Nandi, and Michaela Benzeval, 2020. ‘Understanding Society at Ten Years.’ European Sociological Review 36(6): 976–988. doi: 10.1093/esr/jcaa031.

Präg, Patrick and Alexi Gugushvili, 2021. ‘Subjective Social Mobility and Health in Germany.’ European Societies 23(4): 1–23. doi:10.1080/14616696.2021.1887916.

Präg, Patrick and Lindsay Richards, 2019. ‘Intergenerational Social Mobility and Allostatic Load in Great Britain.’ Journal of Epidemiology and Community Health 73(2): 100–105. doi: 10.1136/jech-2017-210171.

Proto, Eugenio and Climent Quintana-Domeque, 2021. ‘COVID-19 and Mental Health Deterioration by Ethnicity and Gender in the UK.’ Plos One 16(1): e0244419. doi: 10.1371/journal.pone.0244419.

Rose, David and David J. Pevalin, 2003. A Researcher’s Guide to the National Statistics Socio-Economic Classification. London: Sage. doi:10.4135/9780857024725.

Schmid, Lisa, Jonathan Wörn, Karsten Hank, Barbara Sawatzki, and Sabine Walper, 2021. ‘Changes in Employment and Relationship Satisfaction in Times of the COVID-19 Pandemic: Evidence from the German Family Panel.’ European Societies 23(S1): S743–S758. doi:10.1080/14616696.2020.1836385.

Shen, Jing and David Bartram, 2021. ‘Fare Differently, Feel Differently. Mental Wellbeing of UK-Born and Foreign-Born Working Men During the COVID-19 Pandemic.’ European Societies 23(1): S370–S383. doi:10.1080/14616696.2020.1826557.

Thoits, Peggy A., 2010. ‘Stress and Health: Major Findings and Policy Implications.’ Journal of Health and Social Behavior 51(Supplement): S41–S53. doi: 10.1177/0022146510383498.

University of Essex and Institute for Social and Economic Research, 2021. ‘Understanding Society: COVID-19 Study 2020–21.’ doi: 10.5255/UKDA-SN-8644-9.

van der Nest, Gavin, Valéria Lima Passos, Math J. J. M. Candel, and Gerard J. P. van Breukelen, 2020. ‘An Overview of Mixture Modeling for Latent Evolutions in Longitudinal Data: Modeling Approaches, Fit Statistics, and Software.’ Advances in Life Course Research 43(100323): 1–17. doi:10.1016/j.alcr.2019.100323.

van Tilburg, Theo G., Stephanie Steinmetz, Elske Stolte, Henriëtte van der Roest, and Daniel H. de Vries, 2021. ‘Loneliness and Mental Health During the COVID-19 Pandemic: A Study Among Dutch Older Adults.’ Journals of Gerontology B doi: 10.1093/geronb/gbaa111.

Van Winkle, Zachary, Emanuele Ferragina, and Ettore Recchi, forthcoming. ‘The Unexpected Decline in Feelings of Depression Among Adults Aged 50 and Older in Eleven European Countries Amidst the COVID-19 Pandemic.’ Socius 7.

Witteveen, Dirk and Eva Velthorst, 2020. ‘Economic Hardship and Mental Health Complaints during COVID-19.’ Proceedings of the National Academy of Sciences 117(44): 27277–27284. doi:10.1073/pnas.2009609117.

Wright, Liam, Andrew Steptoe, and Daisy Fancourt, 2021. ‘Does Thinking Make It So? Differential Associations Between Adversity Worries and Experiences and Mental Health During the COVID-19 Pandemic.’ Journal of Epidemiology and Community Health pp. 1–7. doi: 10.1136/jech-2020-215598.

Zoch, Gundula, Ann-Christin Bächmann, and Basha Vicari, 2021. ‘Who Cares When Care Closes? Care Arrangements and Parental Working Conditions During the COVID-19 Pandemic in Germany.’ European Societies 23(S1): S576–S588. doi: 10.1080/14616696.2020.1832700.

